# Can Parents and Patients Understand Myopia Using Large Language Model-Based Chatbots?

**DOI:** 10.64898/2026.03.09.26347905

**Authors:** Swati Panigrahi, Sujit Shah, Swapnil Thakur, Sayantan Biswas, Pavan K. Verkicharla

**Affiliations:** Prof. Brien Holden Eye Research Centre, Brien Holden Institute of Optometry and Vision Sciences, LV Prasad Eye Institute, Hyderabad, India, 500034; School of Optometry, College of Health and Life Sciences, Aston University, Birmingham B4 7ET, UK; Infor Myopia Centre, LV Prasad Eye Institute, Hyderabad, India, 500034

**Keywords:** AI chatbot, ChatGPT, DeepSeek, Gemini, myopia

## Abstract

**Purpose:** This study aimed to compare the reliability of myopia-related information from AI chatbots using a set of commonly asked questions by parents and patients on myopia, which is an emerging disease of the 21^st^-century.

**Design:** Prospective comparative reliability study

**Methods:** The study used ChatGPT(OpenAI(2025)GPT-5), Gemini(Gemini 2.0,Google,2025) and DeepSeek (DeepSeek-R1). Twenty myopia-related questions were framed from the perspective of parents and patients, covering general questions, prevention and control, and complications of myopia. Based on their experience in the field of myopia, two senior clinicians, one junior clinician and one researcher(all≥3 years of experience in myopia) rated the responses generated by AI chatbots on a 5-point Likert scale(1:very poor, 2:poor, 3: acceptable, 4:good and 5:very good).

**Results:** Overall, combined rating for tested chatbots had median score of 4(“good”). Gemini received significantly lower ratings than other two chatbots (p≤0.001), with a median rating of 3(“acceptable”). ChatGPT and DeepSeek had median score of 4(“good”) and there was no significant difference in ratings (p=0.48). Both ChatGPT(66.0%) and DeepSeek(67.5%) had high proportions of “good” and “very good” ratings, compared to Gemini(40.0%). Combined “poor” and “very poor” ratings were highest for Gemini(7.5%), followed by ChatGPT(5.0%) and DeepSeek(4.0%). For general questions on myopia, ChatGPT and DeepSeek were rated “good”; for complications of myopia, ChatGPT was rated as “good”, while others were rated “acceptable”.

**Conclusions:** ChatGPT and DeepSeek demonstrated consistently high-quality responses, while ratings for Gemini were slightly lower but remained adequate. These findings suggest AI chatbots can support patients or parents in understanding myopia.

## Introduction

The post-COVID era has seen a significant advancement in Artificial Intelligence (AI), internet accessibility by the global population or to seek spontaneous answers for their health conditions.^[1]^ Patients and their caregivers or family members increasingly rely on the internet and social media for health-related information,^[2]^ but the reliability of the information varies depending on the source.^[3, 4]^ Recently introduced AI chatbots may help bridge this gap by providing evidence-based guidance, but the discrepancies between professional advice and online sources can still be present and create confusion and dissatisfaction among the seekers.^[5]^ ChatGPT (OpenAI), Gemini (Google DeepMind), and DeepSeek (DeepSeek AI) are among the latest large language model-based AI chatbots designed to generate human-like responses and are freely available online. These models have been leveraged with supervised learning strategies and have recently gained widespread attention in the medical community.^[6]^ Although not originally developed for healthcare, the growing popularity and frequent use of these chatbots by the general public (approximately 80-90%)^[7, 8]^ present an opportunity to explore their potential in addressing ophthalmic queries for common ocular diseases. Recently, several studies have assessed the accuracy and reliability of responses provided by artificial intelligence chatbots on ocular conditions, including cataract,^[9]^ glaucoma,^[10]^ and amblyopia.^[11]^

Myopia has become a global concern and is emerging as a public health concern due to an increase in prevalence and its associated vision-threatening ocular complications.^[12]^ The associated personal impact of myopia, such as high economic burden,^[13]^ poor quality of life,^[14]^ and loss of job opportunities^[15]^ further intensifies its societal impact. While awareness about myopia is increasing among clinicians,^[16]^ it remains suboptimal among parents or caregivers.^[17, 18]^ With the increasing awareness initiatives by organisations like Global Myopia Awareness Coalition, individuals with myopia or parents of children with myopia may seek information about myopia through the internet and social media.^[19]^ Given the growing perception of AI chatbots as advanced and easily accessible online search tools, it is important to evaluate the reliability of the information they provide in response to the questions on myopia from parents and patients.

Biswas et al.^[20]^ assessed the accuracy of ChatGPT in answering a series of 11 questions related to myopia. While the findings highlight its potential in rapidly providing information on myopia, the authors also mention some concerns in the chatbot providing inaccurate responses. Given the increasing popularity of various AI chatbots like Gemini and DeepSeek, we aimed to compare the reliability of three AI chatbots (ChatGPT, Gemini, and DeepSeek) in answering the patient and parent-driven questions on myopia and its control. This study concentrated on questions typically raised by patients or parents in real-world clinical settings.^[20]^

## Methods

This cross-sectional study was conducted at the Myopia Research Lab, LV Prasad Eye Institute between February to March 2025. No human participants were recruited for this study; therefore, ethical approval was not required. The study protocol involved three sequential stages (Figure 1): i) determining the number and content of the myopia-related questions, ii) inputting finalised full-text questions into three AI chatbots (ChatGPT (OpenAI. (2025). GPT-5), Gemini (Gemini 2.0, Google, 2025), and DeepSeek (Version DeepSeek V3)) and entering the questions with generated responses in MS-Excel (version 2505) iii) to obtain rating from raters (n=4 raters) including two senior clinicians, one junior clinicians, and one myopia researcher. Senior clinicians have more than 5 years of experience in myopia management, while junior clinician had 3 years of clinical experience in the myopia management. One myopia researcher had 4 years of research experience in myopia. To provide perspective, the myopia clinic provides care to an average of 12 to 15 patients per day, resulting in an annual patient volume of over 10,000.

**Figure 1.**
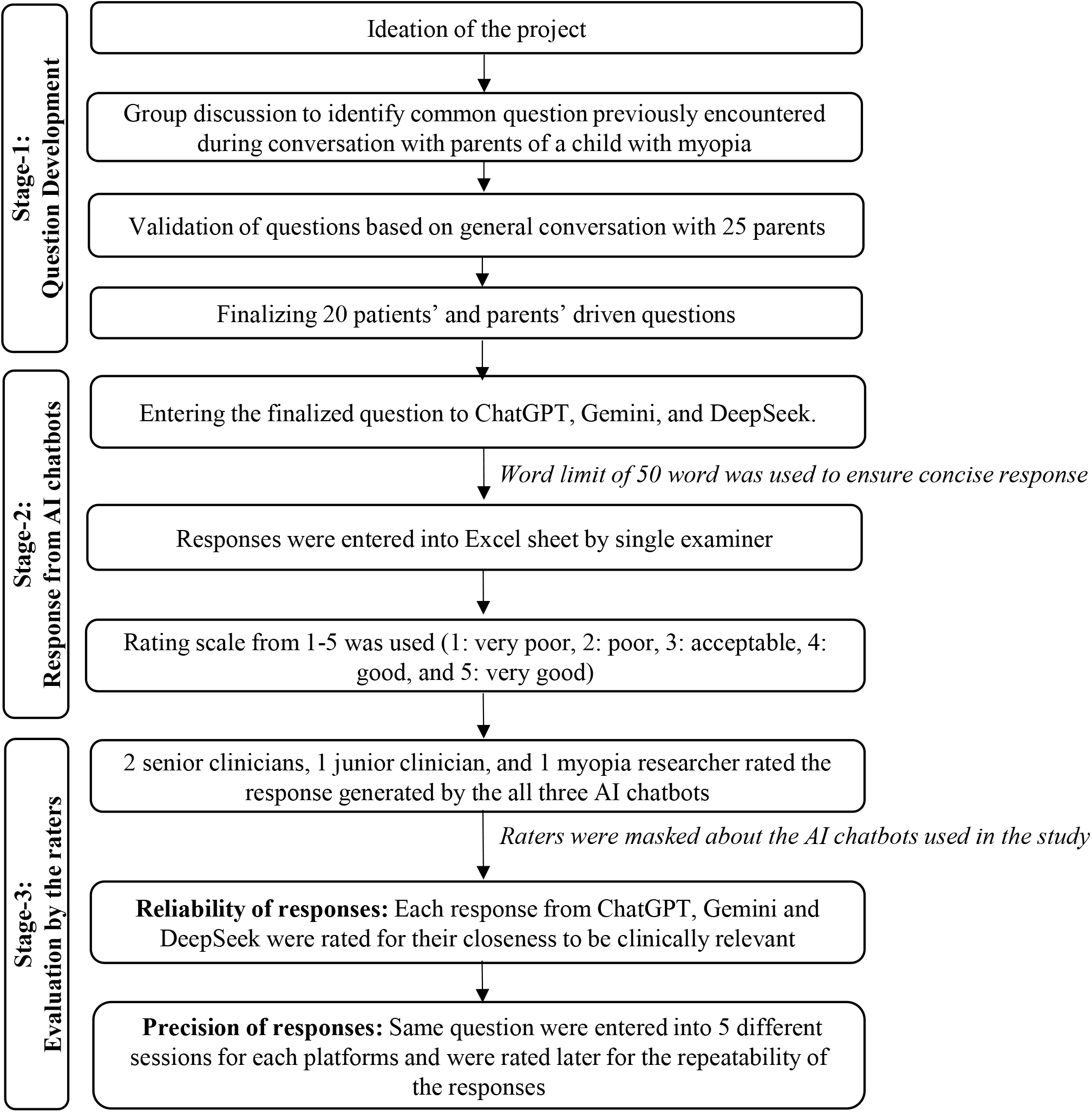
Flowchart of the study methodology illustrating Question development, response generation and evaluation by junior clinician, senior clinician and myopia researcher.

In the first stage, clinicians, researchers, and optometry fellows pursuing myopia fellowship at the Infor Myopia Centre participated in a group discussion to finalise a set of questions informed by the perspectives of patient and parents. Involving a group that regularly engages with patients and parents helped ensure the inclusion of real-world questions that closely reflect those commonly raised by parents in clinical settings. To assess the validity of the questions, one of the examiners (SS) asked 25 parents (whose children has myopia) in a general conversation to share what kind of questions they would ask to AI chatbots to learn about myopia. Based on this conversation, a total of 20 questions were finalised which were categorised into 3 groups: a) general questions on myopia, b) myopia control and prevention and c) complications of myopia (Supplementary Table 1). Question 1 to 8 represents “general questions on myopia”, question 9 to 15 represents “questions related to myopia control and prevention” and question 16 to 20 represents “question related to complications of myopia”. Considering response of patients, there were more general questions on myopia than related to control, prevention and complications of myopia.

In the second stage, the questions were entered one by one into all three AI chatbots— ChatGPT, Gemini, and DeepSeek in a single session. Parents recommended keeping chatbot responses brief and concise to enhance the readability and accessibility of scientific content for laypersons. To ensure consistency in response length, reduce cognitive load, accommodate readers with potential reading difficulties,^[21]^ and improve the accessibility of the scientific content, AI chatbots were instructed to generate responses limited to 50 words. The responses were saved and recorded in an Excel sheet by the same examiner (SP). The MS Excel sheet (version 2505) was then shared with 4 masked raters to assess the reliability of the responses. In the third stage, the raters were asked to evaluate the responses using a 5-point Likert scale, where higher scores indicated greater reliability: 1 – very poor, 2 – poor, 3 – acceptable, 4 – good, 5 – very good. To improve consistency, raters were provided with exemplar responses illustrating each score level, and a brief calibration session was conducted (Table 1). It is worth highlighting that the raters were masked to the type of chatbot used and the excel sheet has headers chatbot-1, 2 and 3. The responses were randomised in the columns (That means the response for Chat GPT may present in column 1, 2 or 3).

**Table 1.**
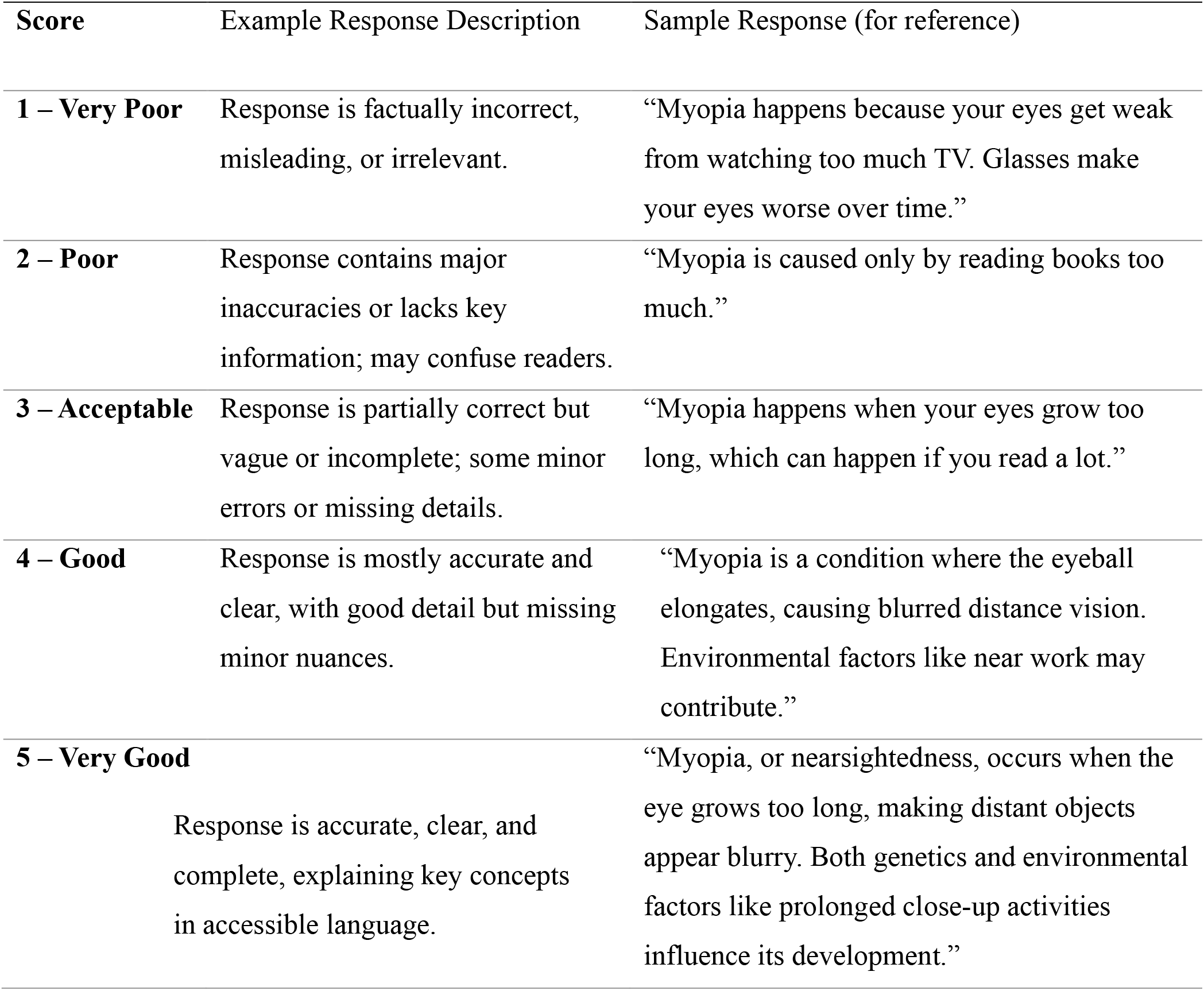
Exemplar responses illustrating each score level.

To evaluate the internal consistency of the response to 20 questions among the three chatbots, Cronbach’s α was calculated. The Cronbach’s α value of 0.7 is considered acceptable.^[22]^ Considering that the AI based models improve itself constantly, we evaluated the repeatability of the responses from each chatbot using a set of 5 randomly selected questions (Question No.2, 4, 10, 13 and 20). For this, the responses from chatbots were obtained by entering the same 5 questions in five different computers repeated on 2 different days. The questions were entered with 5 different login IDs in fresh session for each question to reduce the influence of previous question. To avoid bias, the responses were randomly placed in different columns with headers as 1, 2 and 3 (that means there is possibility that the responses of ChatGPT may present in column 1 or 2 or 3, same was for other chatbots)

To evaluate the intra-rater reliability,^[23]^ 5 randomly selected questions (question no. 1, 3, 9, 12 and 19) with same responses were provided to the raters in 2 different time points with 3 weeks of interval. The intra class correlation coefficient (ICC) was obtained for the ratings between 1^st^ and 2^nd^ time points with two-way mixed effect for consistency and average measures. To avoid bias, the responses were randomly placed in different columns with headers as 1, 2 and 3 (that means there is possibility that the responses of ChatGPT may present in column 1 or 2 or 3, same was for other chat bots). An ICC value of <0.40 considered poor agreement, 0.40-0.59 considered fair agreement, 0.60-0.74 considered good agreement and ≥0.75 considered excellent agreement.^[24]^ Table 3 shows the ratings of 4 raters in two different time points.

To evaluate the inter-rater agreement, intra class correlation coefficient (ICC) was obtained for the ratings between raters with two-way random effect for absolute agreement and average measures for each chatbots.

## Statistical analysis

Data were analysed using IBM SPSS Statistics 26 software (version 26, IBM Corp., Armonk, NY) and graphs were plotted using MS-Excel sheet (version 2505). Rating of responses were reported as median (Q1, Q3) values. To evaluate the internal consistency and repeatability of responses Cronbach’s α was reported. ICC was used to assess inter-rater agreement and intra-rater reliability among the 4 raters. To compare the scores between AI chatbots Kruskal-Wallis H test and Mann-Whitney U test was used. p<0.05 was considered statistically significant.

## Results

Overall, the combined rating i.e. an assessment of rating for the tested chatbots together revealed a median rating of 4.00 (Q1, Q3; 3.00, 4.00) indicating “good” responses. Based on categories of questions, for general questions on myopia, the responses from ChatGPT and DeepSeek were rated as “good”, with a median rating of 4.00 (3.00, 4.00), while Gemini was rated as “acceptable” (3.00, (3.00, 4.00)). For questions related to myopia control and prevention, all the three AI chatbots were rated as good: ChatGPT (4.00 (4.00,4.00)), Gemini (4.00 (3.00, 4.00)) and DeepSeek (4.00 (4.00, 5.00)). For questions related to complications of myopia, the responses of ChatGPT were rated as “good” (4.00 (3.00,4.00)), whereas Gemini (3.00 (3.00, 3.75)) and DeepSeek (3.00 (3.00, 4.00)) were rated as “acceptable”. For independent analysis of ratings for responses from each chatbot, the responses from ChatGPT and DeepSeek were rated as “good” with a score of 4.00 (3.00, 4.00), however, the response from Gemini was rated as “acceptable” (3.00 (3.00,4.00)). The median rating for responses generated by each chatbot is given in Table 2.

**Table 2.** Comparison of median (Q1, Q3) score of individual questions across 3 AI chatbots. ‘*’ represents p<0.05.

| Category of questions |  | ChatGPT (median (Q1, Q3)) | Gemini (median (Q1, Q3)) | DeepSeek (median (Q1, Q3)) | p-value |
| --- | --- | --- | --- | --- | --- |
| 1 | General question on myopia (Q1-Q8) | 4.00 (4.00, 4.00) | 3.00 (3.00, 3.75) | 3.50 (3.00, 4.75) | 0.19 |
| 2 |  | 4.00 (3.25, 4.75) | 3.00 (3.00, 3.75) | 3.50 (3.00, 4.00) | 0.30 |
| 3 |  | 4.00 (3.25, 4.00) | 3.00 (2.25, 3.75) | 4.50 (4.00, 5.00) | 0.04* |
| 4 |  | 3.00 (2.25, 3.75) | 3.00 (2.25, 3.75) | 3.50 (3.00, 4.00) | 0.54 |
| 5 |  | 4.00 (3.25, 4.75) | 4.00 (3.25, 4.00) | 4.00 (3.25, 4.00) | 0.83 |
| 6 |  | 3.00 (2.25, 3.75) | 3.50 (2.25, 4.00) | 4.00 (3.00, 5.00) | 0.42 |
| 7 |  | 3.50 (3.00, 4.00) | 3.00 (3.00, 4.50) | 4.00 (4.00, 4.75) | 0.22 |
| 8 |  | 2.50 (2.00, 3.00) | 3.50 (3.00, 4.00) | 4.00 (3.25, 4.75) | 0.05 |
| 9 | Questions related to myopia control and prevention (Q9-Q15) | 4.00 (3.25, 4.00) | 4.00 (3.25, 4.00) | 4.00 (4.00, 4.75) | 0.29 |
| 10 |  | 4.00 (3.25, 4.75) | 3.50 (3.00, 4.00) | 3.50 (3.00, 4.75) | 0.26 |
| 11 |  | 4.00 (4.00, 4.00) | 3.00 (3.00, 3.75) | 3.50 (3.00, 4.75) | 0.19 |
| 12 |  | 4.00 (4.00, 4.75) | 4.00 (3.25, 4.00) | 4.50 (3.25, 5.00) | 0.46 |

A significant difference was found among three AI chatbots based on the rating for all the responses (Kruskal Wallis test, p< 0.001). Further pair-wise comparison of AI chatbots using Mann-Whitney U test indicated that ratings for responses from Gemini (p≤0.001) were significantly different compared to the other two models, there was no significant difference between the ratings for ChatGPT and DeepSeek (p=0.48).

Except for three responses i.e., question number 3 (general questions on myopia), 18 (complications of myopia) and 20 (complications of myopia), there was no significant difference in median ratings of responses among the three chatbots (Table 2). For question 3, “If my child uses mobile phone at far distance, will he/she get myopia? Give answer in 50 words,” ChatGPT and DeepSeek were rated as “good” with median scores of 4.00 (3.25, 4.00) and 4.50 (4.00, 5.00), respectively, compared to an “average” rating for Gemini with a median score of 3.00 (2.25, 3.75) (p=0.04). For question 18, “Why eye ball size increases? Give answer in 50 words,” ChatGPT was rated as “good” with a median score of 4.00 (4.00, 4.00), compared to “average” ratings for Gemini and DeepSeek with a median score of 3.00 (3.00, 3.00) and 3.00 (3.00, 3,75) (p=0.02). For question 20, ChatGPT and DeepSeek received median scores of 4.00 (3.25, 4.00) and 4.00 (4.00, 4.00) indicating “good” responses, compared to an “acceptable” response from Gemini with a median score of 3.00 (3.00, 3.00) (p=0.01),.

Figures 2 shows the combined ratings of 4 raters for the responses to 20 questions by ChatGPT, Gemini, and DeepSeek. Out of 80 ratings, while ChatGPT and DeepSeek received similar percentage of “very good” and “good” ratings (ChatGPT 66.0%, DeepSeek 67.5%), Gemini had relatively lower proportion of ratings in this category (40.0%). DeepSeek received the highest number of “very good” ratings (19.0%), followed by ChatGPT (12.5%) and Gemini (2.50%). The proportions of ratings for “poor” and “very poor” combined were high for Gemini (7.5%), followed by ChatGPT (5.0%) and DeepSeek (4.0%). Overall “very poor” ratings were rare across all the chatbots (0.0-2.5%). Gemini received the highest number of “acceptable” ratings, with 42.0 (52.5%), compared to 23.0 (29.0%) for ChatGPT and DeepSeek.

**Figure 2.**
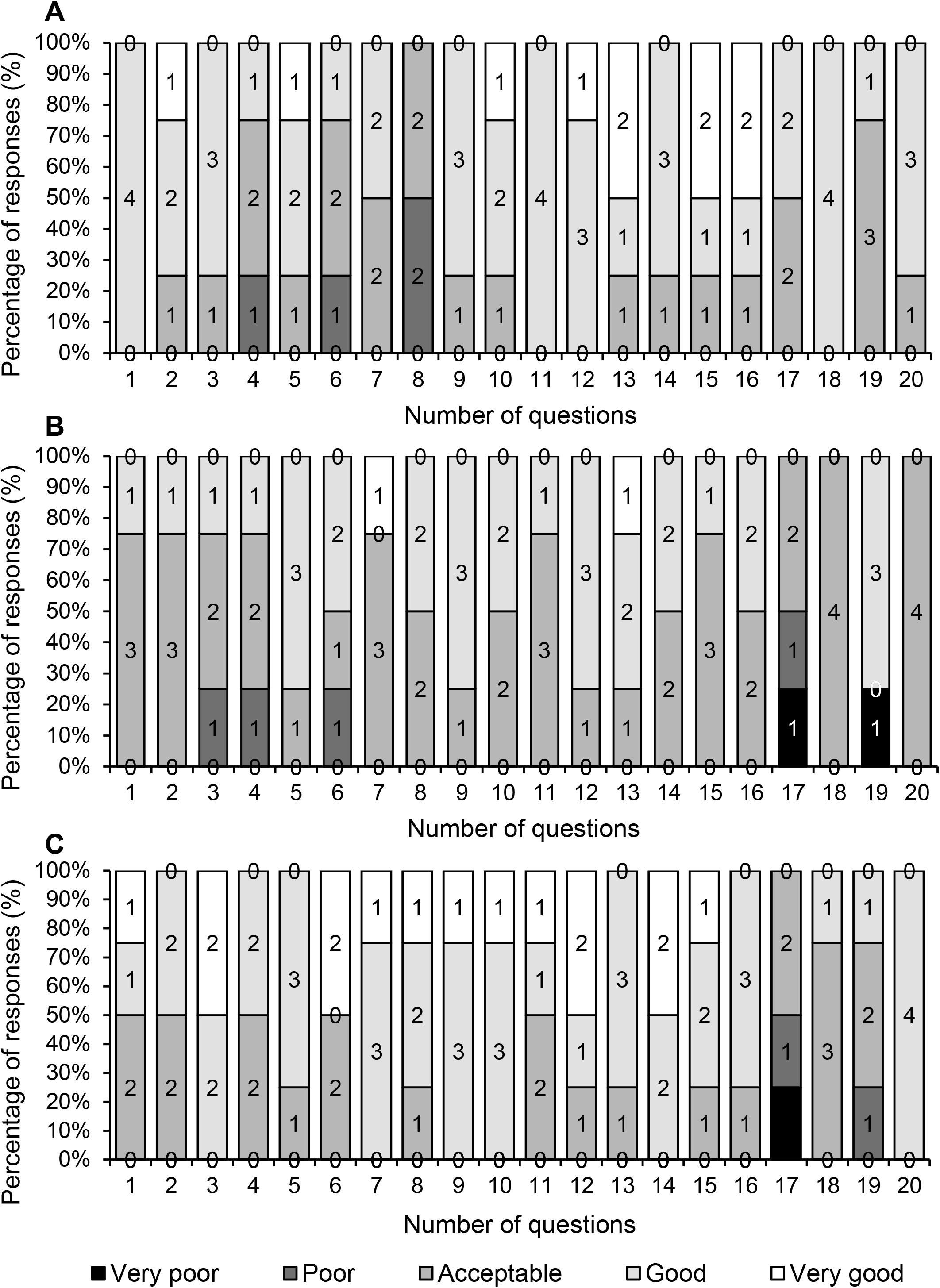
Column chart represents the distribution of combined response of 4 raters for A) ChatGPT, B) Gemini and C) DeepSeek for each question. Numbers on the columns represents the frequency of responses for that specific rating. Black columns: Very poor, Dark Grey columns: Poor, Grey columns: Acceptable, Light Grey columns: Good and White columns: Very Good.

The internal consistency of the 20 responses among the three AI chatbots was analysed based on Cronbach’s α. The Cronbach’s α was 0.70. The Cronbach’s α value of 0.70 is considered acceptable.^[22]^

The repeatability of the responses for same questions from each chatbot in 2 different time points were analysed with Cronbach’s α which revealed the value of -0.26 for ChatGPT, -0.11 for Gemini and 0.37 for DeepSeek. The findings indicate, responses generated by each chatbot were different when prompted again in different time points with different login ID.

For the intra-rater reliability, the ICC values ranged between 0.36 to 0.58, with 0.36 (95% CI: -0.91, 0.78) and 0.41 (95% CI: -0.75, 0.80) for senior clinicians, 0.58 (95% CI: -0.24, 0.86) for junior clinician and 0.45 (95% CI: -0.65, 0.10) for myopia researcher. An ICC value of <0.40 considered poor agreement, 0.40-0.59 considered fair agreement, 0.60-0.74 considered good agreement and ≥0.75 considered excellent agreement.^[22]^ Table 3 shows the ratings of 4 raters in two different time points.

**Table 3.**
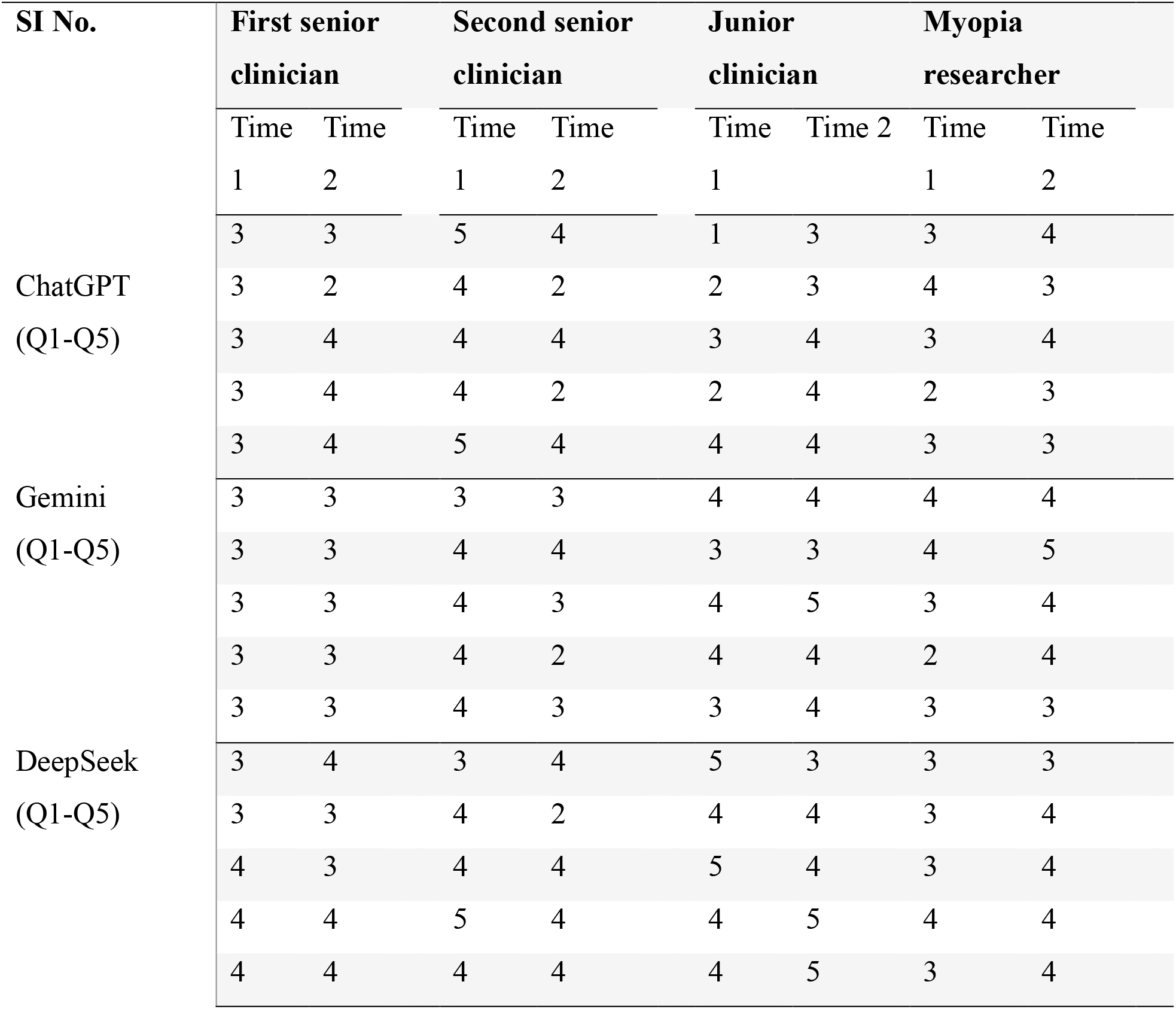
Represents the ratings for the same responses by ChatGPT, Gemini and DeepSeek at two time points.

For inter-rater agreement, the ICC values for ChatGPT was 0.61 (95% CI: 0.25, 0.82), for Gemini 0.13 (95%CI: -0.70, 0.61) and for DeepSeek 0.57 (95% CI: 0.19, 0.81). Table 4 shows the distribution of ratings for three AI chatbots by two senior clinicians, one junior clinician, and one myopia researcher. With respect to inter-rater agreement, for ChatGPT and DeepSeek, out of 4, 3 raters had identical median score of 4.00 (good rating) and for Gemini, 3 raters out of 4 had identical score of 3.00 (acceptable rating).

**Table 4.**
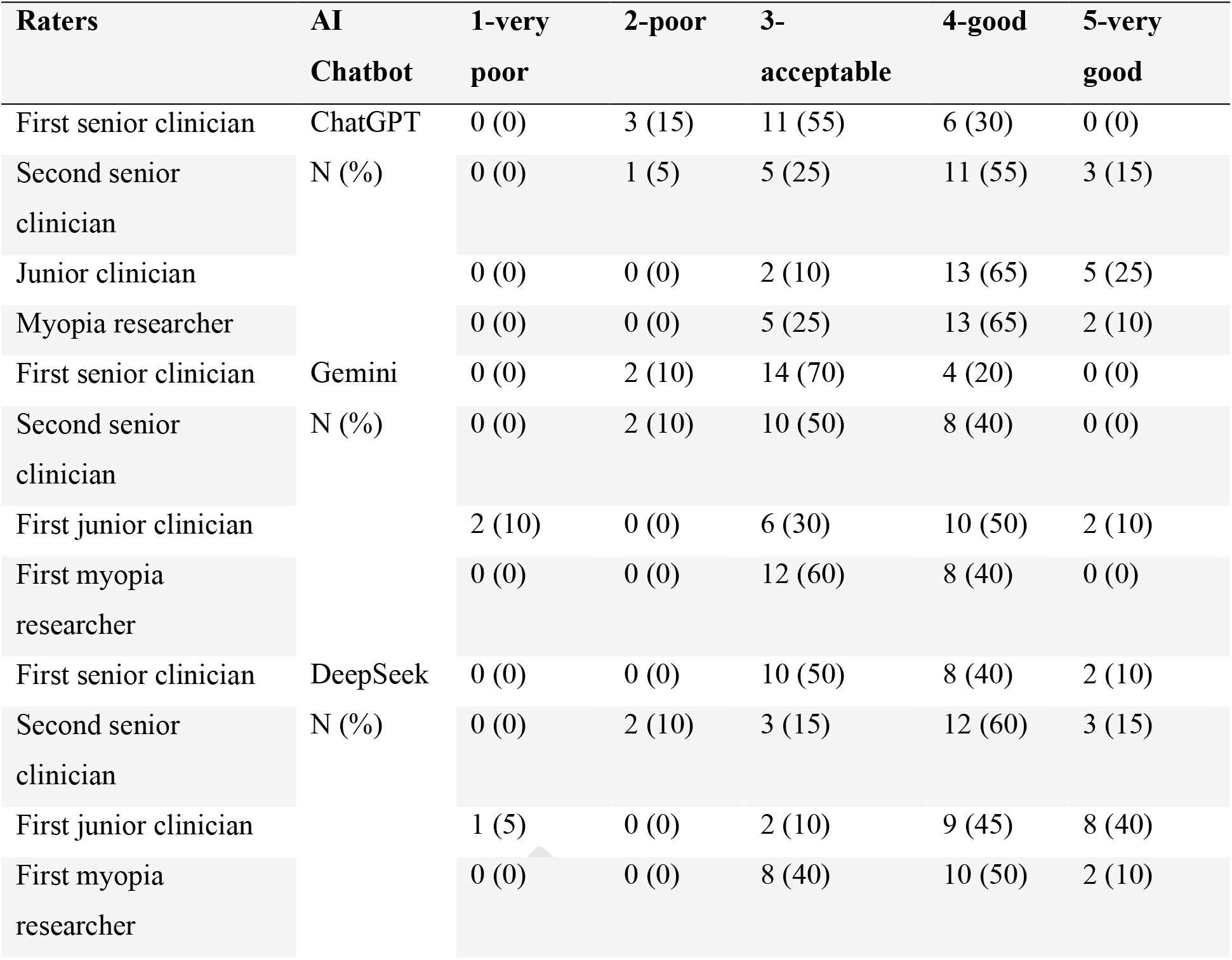
Proportion of ratings for 2 senior clinicians, 2 junior clinicians, and 2 myopia researchers.

## Discussion

AI chatbots such as ChatGPT, DeepSeek, and Gemini are increasingly being explored as tools to address health-related queries for initial patient and caregiver education due to their ease of access, free availability, and capacity to provide quick, conversational responses. However, the reliability of their responses is heavily dependent on the quality of their training data, alignment with current scientific evidence, and ability to understand clinical context. Current study evaluated the ability of three AI chatbots to generate accurate responses related to myopia which were frequently asked by patient and parents. By involving masked raters with different level of experience and generating questions based on parent and patient perspective with focused group discussion this study meticulously reviews the responses of the three AI chatbots.

Three main findings are worth highlighting from the current study. First, overall, the responses generated by AI chatbots were rated as “good” for questions related to myopia. Second, among all the three, DeepSeek (67.5%) and ChaGPT (66.0%) were rated as “good” and “very good”, while Gemini (40.0%) had the lowest performance with fewer “good” and a significant “poor” rating. Third, very few responses from AI chatbots were consistently rated as “very poor” (0.0-2.5%) across all raters.

Similar to previous studies, in the current study the response from ChatGPT was rated as “good” (54.0%). The percentage of “good” responses were ranging from 49.0% to 70.0% in different studies.^[20, 25, 26]^ The differences in percentage values could be due to the AI chatbots are constantly learning and evolving. Furthermore, the inter-rater agreement can be poor among experts. Among the AI models, the ratings for ChatGPT and DeepSeek were similar in the current study, which is in line with findings of Sallam et al.^[27]^ Gemini was rated with higher “poor” ratings compared to ChatGPT in the current study. Delsoz et al. reported that for producing patient education materials related to childhood myopia ChatGPT performed better compared to Gemini.^[28]^

Among different categories of questions, for general questions related to myopia ChatGPT and DeepSeek were rated as “good” while Gemini was rated as “acceptable”. For question related to complication of myopia, ChatGPT performed better than Gemini and DeepSeek. However, for questions on myopia control and prevention all the three AI chatbots rated equally. This is the first study to compare the response of these three AI chatbot in response to questions for above category.

Despite the overall trends, there were few instances such as for question “Is myopia reversible? Give answer in 50 words” ChatGPT provided poor response while Gemini and DeepSeek provided better responses explaining myopia can be controlled and corrected, but is not reversible. Notably, Gemini frequently referred atropine as a treatment option for myopia, but often omitted specifying low-dose atropine, which is the commonly recommended form for myopia control. There was also variability in rating between senior clinicians, researchers and junior clinicians. A study done by Sanchez Tena et al.^[29]^ reported that there is possibility in variable ratings between raters with different level of experience, which could be the reason behind the variability in the inter-rater agreement for the three AI chatbots.

Overall, AI chatbots like ChatGPT, DeepSeek, and Gemini provide quick and accessible responses to patient and parent inquiries, but their reliability depends on the accuracy of their training data and contextual understanding. One of the greatest advantages of these tools are, it is easy to access and free of charge.

This is the first study to compare and evaluate the reliability of ChatGPT, DeepSeek, and Gemini in the context of myopia-related queries based on parent and patient driven questions. However, there are few limitations. The questions posed to the AI chatbots were systematically developed by experienced clinicians, drawing on extensive clinical encounters and documented parent inquiries to closely approximate real-world concerns. While these questions may not fully capture the diversity of real-world queries posed to AI chatbots, the structured development process aimed to ensure they were representative and relevant for this study. Responses were limited to a 50-word limit. While this was strategically done uniformly across all models to maintain comparability, to minimise the cognitive load while reading and accessibility of the scientific content, the word limit may have restricted the depth, nuance and completeness of the responses.^[30]^

In conclusion, ChatGPT and DeepSeek demonstrated comparable performance, delivering consistently high-quality responses. In contrast, output of Gemini was slightly lower but remained adequate. Findings suggest that AI chatbots could support patient or parents in improving their understanding of myopia and enhancing their awareness. Enhancing these tools with evidence-based training, contextual understanding, and multilingual support could significantly improve their utility for public health communication and patient guidance in ophthalmology and beyond.

## Data Availability

All data produced in the present work are contained in the manuscript

## Acknowledgments

The authors acknowledge Hyderabad Eye Research Foundation, Hyderabad, India for their support. Authors also acknowledge the myopia fellows, researchers and clinicians for their support with question generation.

## References

1. Hussain T, Wang D, Li B. The influence of the COVID-19 pandemic on the adoption and impact of AI ChatGPT: Challenges, applications, and ethical considerations. Acta Psychol (Amst). 2024;246:104264.

2. Van de Belt TH, Engelen LJ, Berben SA, Teerenstra S, Samsom M, Schoonhoven L. Internet and social media for health-related information and communication in health care: preferences of the Dutch general population. J Med Internet Res. 2013;15(10):e220.

3. Goodman RS, Patrinely JR, Stone CA, Jr., Zimmerman E, Donald RR, Chang SS, et al. Accuracy and Reliability of Chatbot Responses to Physician Questions. JAMA Netw Open. 2023;6(10):e2336483.

4. Dalmer NK. Questioning reliability assessments of health information on social media. J Med Libr Assoc. 2017;105(1):61–8.

5. Meyrowitsch DW, Jensen AK, Sørensen JB, Varga TV. AI chatbots and (mis)information in public health: impact on vulnerable communities. Front Public Health. 2023;11:1226776.

6. Potapenko I, Boberg-Ans LC, Stormly Hansen M, Klefter ON, van Dijk EHC, Subhi Y. Artificial intelligence-based chatbot patient information on common retinal diseases using ChatGPT. Acta Ophthalmol. 2023;101(7):829–31.

7. Tao W, Yang J, Qu X. Utilization of, Perceptions on, and Intention to Use AI Chatbots Among Medical Students in China: National Cross-Sectional Study. JMIR Med Educ. 2024;10:e57132.

8. Ayo-Ajibola O, Davis RJ, Lin ME, Riddell J, Kravitz RL. Characterizing the Adoption and Experiences of Users of Artificial Intelligence-Generated Health Information in the United States: Cross-Sectional Questionnaire Study. J Med Internet Res. 2024;26:e55138.

9. Cohen SA, Brant A, Fisher AC, Pershing S, Do D, Pan C. Dr. Google vs. Dr. ChatGPT: Exploring the Use of Artificial Intelligence in Ophthalmology by Comparing the Accuracy, Safety, and Readability of Responses to Frequently Asked Patient Questions Regarding Cataracts and Cataract Surgery. Semin Ophthalmol. 2024;39(6):472–9.

10. Wang H, Masselos K, Tong J, Connor HRM, Scully J, Zhang S, et al. ChatGPT for Addressing Patient-centered Frequently Asked Questions in Glaucoma Clinical Practice. Ophthalmol Glaucoma. 2025;8(2):157–66.

11. Nikdel M, Ghadimi H, Tavakoli M, Suh DW. Assessment of the Responses of the Artificial Intelligence-based Chatbot ChatGPT-4 to Frequently Asked Questions About Amblyopia and Childhood Myopia. J Pediatr Ophthalmol Strabismus. 2024;61(2):86–9.

12. Holden BA, Wilson DA, Jong M, Sankaridurg P, Fricke TR, Smith EL, III, et al. Myopia: a growing global problem with sight-threatening complications. Community Eye Health. 2015;28(90):35.

13. Ma Y, Wen Y, Zhong H, Lin S, Liang L, Yang Y, et al. Healthcare utilization and economic burden of myopia in urban China: A nationwide cost-of-illness study. J Glob Health. 2022;12:11003.

14. Wu W, Yi L, Zhang K, Chen Z, Shi C, Chen C, et al. Health-related quality of life measurements in children and adolescents with refractive errors: A scoping review. Adv Ophthalmol Pract Res. 2024;4(2):84–94.

15. Verkicharla PK, Maddali S. Does being a myope reduce opportunities in the Indian armed forces? Indian J Ophthalmol. 2022;70(12):4463–5.

16. Wolffsohn JS, Whayeb Y, Logan NS, Weng R. IMI-Global Trends in Myopia Management Attitudes and Strategies in Clinical Practice-2022 Update. Invest Ophthalmol Vis Sci. 2023;64(6):6.

17. Tian Y, Yu Y. Knowledge, attitude and practice towards myopia among parents of primary school students: a cross-sectional study. BMJ Open. 2025;15(3):e093565.

18. Fu T, Yang Z, Zhang P, Yang J, Gao L. Knowledge, attitude and practice among parents of children and teenagers towards myopia prevention and control during the COVID-19 epidemic. BMJ Open. 2025;15(4):e089431.

19. Ortiz-Peregrina S, Solano-Molina S, Martino F, Castro-Torres JJ, Jiménez JR. Parental awareness of the implications of myopia and strategies to control its progression: A survey-based study. Ophthalmic Physiol Opt. 2023;43(5):1145–59.

20. Biswas S, Logan NS, Davies LN, Sheppard AL, Wolffsohn JS. Assessing the utility of ChatGPT as an artificial intelligence-based large language model for information to answer questions on myopia. Ophthalmic Physiol Opt. 2023;43(6):1562–70.

21. Barton JJ, Hanif HM, Eklinder Björnström L, Hills C. The word-length effect in reading: a review. Cogn Neuropsychol. 2014;31(5-6):378–412.

22. Taber KS. The Use of Cronbach’s Alpha When Developing and Reporting Research Instruments in Science Education. Research in Science Education. 2018;48(6):1273–96.

23. Berchtold A. Test–retest: Agreement or reliability? 2016;9:2059799116672875.

24. Landis JR, Koch GG. The measurement of observer agreement for categorical data. Biometrics. 1977;33(1):159–74.

25. Wang Y, Liang L, Li R, Wang Y, Hao C. Comparison of the Performance of ChatGPT, Claude and Bard in Support of Myopia Prevention and Control. J Multidiscip Healthc. 2024;17:3917–29.

26. Lim ZW, Pushpanathan K, Yew SME, Lai Y, Sun CH, Lam JSH, et al. Benchmarking large language models’ performances for myopia care: a comparative analysis of ChatGPT-3.5, ChatGPT-4.0, and Google Bard. EBioMedicine. 2023;95:104770.

27. Sallam M, Alasfoor IM, Khalid SW, Al-Mulla RI, Al-Farajat A, Mijwil MM, et al. Chinese generative AI models (DeepSeek and Qwen) rival ChatGPT-4 in ophthalmology queries with excellent performance in Arabic and English. Narra J. 2025;5(1):e2371.

28. Delsoz M, Hassan A, Nabavi A, Rahdar A, Fowler B, Kerr NC, et al. Large Language Models: Pioneering New Educational Frontiers in Childhood Myopia. Ophthalmol Ther. 2025;14(6):1281–95.

29. Sanchez Tena MA, Alvarez-Peregrina C, Martinez-Perez C. Evaluation of the perception of information from ChatGPT in myopia education: Perspectives of students and professionals. Ophthalmic Physiol Opt. 2025;45(3):883–94.

30. Pushpanathan K, Lim ZW, Er Yew SM, Chen DZ, Hui’En Lin HA, Lin Goh JH, et al. Popular large language model chatbots’ accuracy, comprehensiveness, and self-awareness in answering ocular symptom queries. iScience. 2023;26(11):108163.

