## Supplementary Table 1 for "Can Parents and Patients Understand Myopia Using Large Language Model-Based Chatbots?"

**Supplementary Data**

**Supplementary Table 1.** Represents the responses obtained from ChatGPT, Gemini and DeepSeek

| SI No. | Categories | Questions | ChatGPT | Gemini | DeepSeek |
| --- | --- | --- | --- | --- | --- |
| 1 | General questions on myopia | What is myopia? Give the answer in 50 words. | Myopia, or nearsightedness, is a common eye condition where distant objects appear blurry while nearby objects remain clear. It occurs when the eyeball is too long or the cornea is too curved, causing light to focus in front of the retina instead of directly on it. Spectacles or contact lenses correct it. | Myopia, also known as nearsightedness, is a common vision condition where you can see objects near to you clearly, but objects farther away appear blurry. It occurs when the eye's shape causes light to bend incorrectly, focusing in front of the retina instead of on it. | Myopia, or nearsightedness, is a common vision condition where distant objects appear blurry while close ones remain clear. It occurs when the eyeball is too long or the cornea is too curved, causing light to focus in front of the retina instead of on it. Corrected with glasses, contacts, or surgery. |
| 2 |  | Can my child wear cap or hold umbrella, while going outdoors to prevent myopia? Give answer in 50 words. | Yes, your child can wear a cap or hold an umbrella while outdoors to protect against harsh sunlight. However, it's essential that they still receive adequate exposure to natural light, as UV protection doesn't block the beneficial wavelengths thought to help prevent myopia. Balancing protection and exposure is key. | While caps and umbrellas protect from sun, they don't directly prevent myopia. The benefit comes from *outdoor time* itself, not just being in the sun. So, yes, your child can wear them, but the key is still spending sufficient time outdoors, even in less sunny conditions. | Yes, your child can wear a cap or hold an umbrella outdoors, as these provide sun protection without significantly reducing the benefits of natural light exposure, which helps prevent myopia. Ensure they still spend 1-2 hours outside daily, as the key factor is overall light exposure, not direct sunlight. |
| 3 |  | If my child uses mobile phone at far distance, will he/she get myopia? Give answer in 50 words. | Using a mobile phone from a far distance reduces the risk of myopia compared to close usage. However, prolonged screen time can still cause digital eye strain and disrupt healthy visual habits. Encourage regular breaks, maintain a balanced screen time, and prioritize outdoor activities to protect your child's eye health. | While using a phone at a distance is better than close-up, excessive screen time, even at a distance, can still contribute to eye strain and potentially increase the risk of myopia development over time. Outdoor time remains crucial. | Using a mobile phone at a far distance reduces eye strain compared to close-up use, but prolonged screen time can still contribute to myopia risk. Encourage breaks, outdoor activities, and the 20-20-20 rule to minimize strain. Balancing screen use with healthy visual habits is key to protecting your child's eyes. |
| 4 |  | Can my child do near work activities during outdoors, for preventing myopia? Give answer in 50 words. | While outdoors, near-work activities like reading or using devices are less effective in preventing myopia. The key benefit of outdoor time is distance-focused visual engagement. Encourage activities like playing, exploring nature, or sports to fully utilize outdoor exposure for eye health and reduce prolonged near-focus, which contributes to myopia. | No, near work outdoors doesn't offer the same protective benefits against myopia as general outdoor activity. The key is the type of light exposure received during outdoor time, which is different from focused near work, even if done outside. | While outdoors, encourage your child to focus on distant views rather than near work activities like reading or using screens. Near work can strain the eyes, counteracting the benefits of outdoor time. Prioritize activities like playing, walking, or sports to maximize natural light exposure and reduce myopia risk effectively. |
| 5 |  | What food shall I give to my child to prevent myopia? Give answer in 50 words. | Provide your child with a diet rich in eye-healthy nutrients to help prevent myopia. Include green leafy vegetables, carrots, citrus fruits, and colorful berries for vitamins A and C. Add omega-3 sources like fish and walnuts for retinal health. Eggs, dairy, and nuts are also beneficial for strong vision. | A diet rich in fruits, vegetables (especially leafy greens), and omega-3 fatty acids supports overall eye health. Foods like carrots, spinach, and fish are good choices. However, diet alone won't prevent myopia; outdoor time is crucial too. | To support eye health and prevent myopia, include foods rich in **vitamin A** (carrots, sweet potatoes), **omega-3 fatty acids** (fish, walnuts), **lutein and zeaxanthin** (leafy greens, eggs), and **vitamin C** (citrus fruits, bell peppers). A balanced diet with these nutrients promotes healthy vision and overall eye development. |
| 6 |  | What causes myopia? Give the answer in 50 words. | Myopia occurs when the eyeball grows too long or the cornea curves too steeply, causing light to focus in front of the retina. Genetic factors, excessive near work, prolonged screen time, and limited outdoor exposure contribute to its development. Early lifestyle interventions can help reduce the risk of progression. | Myopia develops when the eye's shape causes light to focus *in front* of the retina, instead of directly on it. This can be due to the eyeball being too long or the cornea being too curved. Genetics and environmental factors both play a role. | Myopia is caused by a combination of genetic and environmental factors. Genetics play a role if parents are nearsighted. Environmental factors include excessive near work (reading, screens), limited outdoor time, and poor lighting. These factors can lead to elongated eyeballs or overly curved corneas, causing light to focus in front of the retina. |
| 7 |  | Does non-myopic sibling require myopia treatment when they don't have myopia? Give answer in 50 words. | Non-myopic siblings do not require myopia treatment but benefit from preventive strategies like regular outdoor activities and balanced screen time. Since genetics and environment influence myopia, proactive measures help reduce their risk. Regular eye check-ups are essential to detect early signs and maintain healthy visual development. | A non-myopic sibling doesn't need myopia *treatment* if they don't have myopia. However, they should still follow myopia *prevention* strategies, like spending time outdoors and managing near work, as they are still at risk of developing it. | No, a non-myopic sibling does not require myopia treatment if they have no vision issues. However, they should follow preventive measures like spending time outdoors, limiting screen use, and having regular eye check-ups to reduce the risk of developing myopia, especially if there’s a family history of the condition. |
| 8 |  | Is myopia reversible? Give answer in 50 words. | Myopia is not reversible, as it involves permanent elongation of the eyeball. However, its progression can be slowed using treatments like low-dose atropine, orthokeratology, and specialized lenses. Lifestyle changes, such as more outdoor time and limited screen use, help manage myopia but do not reverse existing vision changes. | Myopia, or nearsightedness, is not reversible once the eye has grown longer or the cornea has changed shape. ^1^ However, its progression, especially in children, can be slowed down with treatments like specialized contact lenses, atropine eye drops, and increased outdoor time. These methods aim to manage the elongation of the eye. | No, myopia is not reversible, but its progression can be slowed. Treatments like **orthokeratology**, **low-dose atropine**, and **multifocal lenses** help manage it. **Laser surgery (LASIK/PRK)** can correct vision but doesn’t cure myopia. Early intervention in children is key to preventing high myopia and related complications. |
| 9 | Questions related to myopia control and prevention | How can we prevent myopia? Give answer in 50 words. | Myopia can be prevented by encouraging outdoor activities, as natural light helps slow eye growth. Limit screen time and close-up tasks, ensuring regular breaks. Maintain proper lighting while reading. Use myopia control interventions like specialized contact lenses, spectacles, or low-dose atropine under medical guidance. Regular eye check-ups are essential. | While there's no guaranteed way to *prevent* myopia, some strategies may help slow its progression. These include spending more time outdoors, reducing near work (like reading or screen time), and ensuring adequate light when doing close-up tasks. Regular eye exams are also crucial. | To prevent myopia, spend more time outdoors in natural light, limit screen time, and follow the 20-20-20 rule (every 20 minutes, look at something 20 feet away for 20 seconds). Maintain proper reading distance, ensure good lighting, and have regular eye check-ups. Early detection and healthy habits can slow its progression. |
| 10 |  | How many hours my child should spend in outdoors? Give answer in 50 words. | Children should spend at least 2 to 3 hours outdoors daily to help prevent myopia. Exposure to natural light and engaging in distance-focused activities support healthy eye development. Outdoor play also encourages physical activity, reducing screen time and near work, which are risk factors for myopia progression. | Spending at least 2 hours outdoors daily can help reduce the risk of myopia in children. | Children should spend at least **1-2 hours daily outdoors** to help reduce the risk of myopia. Natural light and focusing on distant objects support healthy eye development. Balancing outdoor activities with limited screen time and proper reading habits can significantly contribute to maintaining good vision and overall eye health. |
| 11 |  | What is the treatment to control myopia? Give answer in 50 words. | Myopia control treatments include low-dose atropine eye drops, orthokeratology (overnight lenses), multifocal contact lenses, and specially designed multifocal or DIMS spectacle lenses. Increasing outdoor time and reducing near work also help. These methods slow eye growth, reducing progression and risk of high myopia-related complications. Regular eye exams are essential. | Myopia control aims to slow down the progression of nearsightedness, especially in children. ^1^ Effective treatment include specialized contact lenses like MiSight or Ortho-K, atropine eye drops, and spending more time outdoors. These methods can help manage eye growth and reduce the risk of high myopia later in life. | To control myopia, use **orthokeratology (Ortho-K) lenses**, **low-dose atropine eye drops**, or **multifocal glasses/contact lenses**. Increase **outdoor time** (2+ hours daily) and practice **good visual habits** (20-20-20 rule). Regular **eye exams** are essential. Early intervention slows progression, especially in children. Consult an eye specialist for personalized treatment. |
| 12 |  | Up to how many years, my child should undergo myopia control and myopia prevention strategies? Give answer in 50 words. | Myopia control and prevention strategies should continue until your child's eye growth stabilizes, typically around 16 to 18 years. However, this may vary. Regular eye check-ups are essential to monitor progression and adjust interventions. Early and consistent management reduces the risk of high myopia and associated eye complications later in life. | Myopia control and prevention strategies are generally recommended throughout childhood and adolescence, typically until the late teens or early twenties when eye growth slows down significantly. | Myopia control and prevention strategies should be followed until your child reaches **early adulthood (around 20-21 years)**, when eye growth typically stabilizes. Regular eye check-ups and consistent habits like outdoor time, limited screen use, and proper lighting are essential during childhood and teenage years to manage myopia progression effectively. |
| 13 |  | At what time my child should have sunlight exposure? Give answer in 50 words. | The best time for children to get sunlight exposure is between 7 AM and 10 AM when UV levels are lower and light is safe yet beneficial. This exposure supports healthy eye development and helps reduce myopia risk. Avoid intense midday sun to protect the skin and eyes from UV damage. | Morning or late afternoon sunlight is best for children's eye health. Avoid midday sun due to higher UV risk. Even on cloudy days, outdoor time is beneficial. Always use sun protection. | Your child should have sunlight exposure during **morning (8-10 AM) or late afternoon (4-6 PM)** when UV rays are milder. This ensures safe, beneficial natural light for eye health, reducing myopia risk. Aim for **1-2 hours daily** outdoors, balancing sun protection (caps, sunscreen) with the benefits of sunlight for vision development. |
| 14 |  | Whether eye exercise can help us in the prevention and control of myopia? Give the answer in 50 words. | Eye exercises may help reduce eye strain and improve focusing flexibility but have limited evidence for preventing or controlling myopia. They can complement other measures, such as outdoor activities, reduced screen time, and proper visual habits. Comprehensive myopia management typically involves lifestyle changes and professional interventions when needed. | While eye exercises might reduce eye strain, current scientific evidence doesn't support their effectiveness in preventing or controlling myopia progression. Outdoor time and managing near work are more effective strategies. | Eye exercises may help reduce eye strain but have limited evidence in preventing or controlling myopia. They don’t address the structural causes of myopia, like elongated eyeballs. Combining outdoor activities, reduced screen time, and proper visual habits is more effective for myopia prevention and management. Consult an eye specialist for tailored advice. |
| 15 |  | Which treatment is best for myopia prevention and myopia control? Give answer in 50 words. | The best treatments for myopia prevention and control include lifestyle changes like increasing outdoor time and limiting screen use. Effective clinical interventions are low-dose atropine eye drops, orthokeratology (overnight lenses), and specialized myopia control glasses or contact lenses. Treatment should be personalized based on age, myopia progression, and eye health. | No single "best" treatment exists. For prevention, maximizing outdoor time is key. For control (slowing progression), options include atropine eye drops, multifocal contact lenses, and orthokeratology. An eye doctor can determine the most suitable approach | The best treatments for myopia prevention include increased outdoor time (1-2 hours daily) and reduced near work. For myopia control, options include orthokeratology (Ortho-K) lenses, low-dose atropine eye drops, and multifocal contact lenses or glasses. Consult an eye specialist to determine the most suitable approach for your child. |
| 16 | Question related to complications of myopia | I have myopia, will my child get myopia? Give the answer in 50 words. | If you have myopia, your child has a higher risk due to genetic factors. However, environmental influences also play a significant role. Encourage outdoor activities, limit screen time, and promote balanced visual habits to reduce the risk. Regular eye check-ups can help detect and manage early signs effectively. | Having myopia does increase your child's risk of developing it, as genetics plays a role. However, it's not guaranteed. Lifestyle factors like spending time outdoors also significantly influence myopia development. | If you have myopia, your child has a higher risk of developing it due to genetic factors. However, environmental factors like outdoor time, limited screen use, and proper lighting also play a role. Encourage healthy visual habits and regular eye check-ups to reduce their risk or manage myopia effectively. |
| 17 |  | I have hyperopia will my child get myopia? Give answer in 50 words. | Having hyperopia does not directly mean your child will develop myopia, but it doesn't eliminate the risk either. Genetics, environmental factors, and lifestyle habits play crucial roles. Encourage regular outdoor activities, balanced screen time, and routine eye check-ups to support your child's eye health and monitor any vision changes. | Having hyperopia (farsightedness) does *not* increase your child's risk of developing myopia (nearsightedness). These are distinct refractive errors. However, your child could still develop myopia independently, as genetics and environmental factors play a role. | Having hyperopia (farsightedness) does not directly increase your child’s risk of myopia. However, myopia risk depends on genetic and environmental factors. Encourage outdoor activities, limit screen time, and ensure regular eye check-ups to reduce their risk. Family history of myopia in other relatives can also influence their likelihood. |
| 18 |  | Why eye ball size increases? Give answer in 50 words. | Eyeball size increases mainly due to excessive growth of the eye’s axial length, often caused by genetics, prolonged near work, and limited outdoor activity. This elongation leads to myopia, where light focuses in front of the retina. Environmental and visual habits during childhood strongly influence this abnormal eye growth. | The eyeball size primarily increases due to a condition called **axial myopia**. This happens when the eyeball grows longer than normal during childhood development. As the eye elongates, the distance between the lens and the retina increases, causing light to focus in front of the retina instead of directly on it. This results in blurry distance vision, which is the hallmark of myopia. | The eyeball elongates (axial myopia) primarily due to **genetic factors** and **excessive near work** (e.g., screens, reading) with **insufficient outdoor light exposure**. This stretches the retina, causing blurred distance vision. **Dopamine regulation** (influenced by sunlight) and **eye growth signals** play a role. Controlling myopia early can slow elongation. |
| 19 |  | If the myopia is not prevented, what are the complications? Give the answer in 50 words. | Uncontrolled myopia increases the risk of serious eye complications, including retinal detachment, glaucoma, cataracts, and myopic maculopathy, which can lead to permanent vision loss. High myopia also affects daily life due to dependence on corrective lenses. Early prevention and control are crucial to protect long-term eye health and visual function. | Uncontrolled myopia increases the risk of serious eye problems later in life, including retinal detachment, glaucoma, cataracts, and myopic maculopathy. These conditions can lead to vision loss and even blindness. | If myopia is not prevented or controlled, it can lead to **high myopia**, increasing the risk of serious complications like **retinal detachment**, **glaucoma**, **cataracts**, and **myopic maculopathy**. These conditions can cause permanent vision loss or impairment, emphasizing the importance of early prevention and regular eye care to protect long-term eye health. |
| 20 |  | Will my child become blind due to myopia? Give answer in 50 words. | Most children with myopia will not go blind, especially with proper care. However, high myopia increases the risk of serious eye conditions like retinal detachment or macular degeneration later in life. Early detection, regular eye exams, and myopia control treatments can help protect your child's vision and reduce long-term risks. | While myopia itself doesn't directly cause blindness, high myopia increases the risk of developing eye conditions that can lead to vision loss, such as retinal detachment, glaucoma, cataracts, and myopic macular degeneration. | No, myopia alone rarely causes blindness. However, **high myopia** (above -6.00D) increases risks of **retinal detachment, glaucoma, or macular degeneration** later in life. Proper management—**regular eye exams, myopia control treatments (atropine, Ortho-K), and healthy habits (outdoor time, screen breaks)**—can significantly reduce risks. Early intervention is key. |
